# A whole food, plant-based diet reduces amino acid levels in patients with metastatic breast cancer

**DOI:** 10.1101/2024.10.09.24315165

**Authors:** TashJaé Q. Scales, Bradley Smith, Lisa M. Blanchard, Nellie Wixom, Emily T. Tuttle, Brian J. Altman, Luke J. Peppone, Joshua Munger, Thomas M. Campbell, Erin K. Campbell, Isaac S. Harris

## Abstract

**Background:** Amino acids are critical to tumor survival. Tumors can acquire amino acids from the surrounding microenvironment, including the serum. Limiting dietary amino acids is suggested to influence their serum levels. Further, a plant-based diet is reported to contain fewer amino acids than an animal-based diet. The extent to which a plant-based diet lowers the serum levels of amino acids in patients with cancer is unclear.

**Methods:** Patients with metastatic breast cancer (n=17) were enrolled in a clinical trial with an ad libitum whole food, plant-based diet for 8 weeks without calorie or portion restriction. Dietary changes by participants were monitored using a three-day food record. Serum was collected from participants at baseline and 8 weeks. Food records and serum were analyzed for metabolic changes.

**Results:** We found that a whole food, plant-based diet resulted in a lower intake of calories, fat, and amino acids and higher levels of fiber. Additionally, body weight, serum insulin, and IGF were reduced in participants. The diet contained lower levels of essential and non-essential amino acids, except for arginine (glutamine and asparagine were not measured). Importantly, the lowered dietary intake of amino acids translated to reduced serum levels of amino acids in participants (5/9 essential amino acids; 4/11 non-essential amino acids).

**Conclusions:** These findings provide a tractable approach to limiting amino acid levels in persons with cancer. This data lays a foundation for studying the relationship between amino acids in patients and tumor progression. Further, a whole-food, plant-based diet has the potential to synergize with cancer therapies that exploit metabolic vulnerabilities.

**Trial Registration:** The clinical trial was registered with ClinicalTrials.gov identifier NCT03045289 on 2017-02-07.

## Background

To grow and survive, cancer cells face increased metabolic demands, including a heightened need for amino acids(1, 2). Tumors obtain amino acids through either intracellular production or extracellular uptake. The expression and activity of key enzymes associated with amino acid production are elevated in tumors compared with non-tumorigenic tissue(3, 4). While numerous efforts have explored blocking these enzymes for a therapeutic benefit to cancer patients(5–9), they have met minimal clinical success. A potential explanation for the lack of efficacy of these drugs in clinical settings is the compensatory uptake of circulating amino acids by tumors. An alternative strategy is to lower the levels of amino acids in circulation(10–13), thus limiting the tumor’s ability to uptake amino acids. Indeed, decreasing the dietary intake of either total protein(14–17) or specific amino acids(18–21) significantly perturbs tumor growth and progression in pre-clinical models of cancer. Although consumption of a healthy diet correlates with lower cancer risk(22), there is a lack of knowledge of how diet impacts amino acid levels in patients with cancer. Even more, it is unclear whether adopting a diet with reduced amino acids is feasible for patients with cancer.

A plant-based diet lower in processed foods has several potential anti-cancer benefits compared to a Western diet (i.e., an animal-based diet higher in processed foods). Higher fiber intake, commonly associated with a plant-based diet, improves a tumor’s response to immunotherapy(23). A plant-based diet also contains less fats, which are directly associated with increased tumor incidence and progression(24–26). Additionally, a low-fat plant-based diet is associated with lower energy intake than a low-carbohydrate animal-based diet(27), suggesting a plant-based diet could potentially limit energy sources available to tumors. Finally, a plant-based diet contains less protein than animal-based diets(28, 29), suggesting a potential approach to deprive tumors of amino acids. However, whether the implementation of a plant-based diet can lower amino acid levels in patients with cancer is largely unknown.

In this study, a whole food, plant-based diet was adhered to by patients with metastatic breast cancer. While this was an ad-libitum diet with no restriction on food consumption, we found that this diet lowered the intake of calories, fat, and total protein in participants. Further, we showed reduced serum insulin and IGF levels, suggesting an approach to limiting pro-tumorigenic signals. Finally, we found that participants who consumed a whole food, plant-based diet had lower levels of amino acids in their serum. Together, these findings suggest that adopting a whole food, plant-based diet is a tractable approach towards lowering circulating amino acid levels in patients with cancer and potentially depleting cancers of key metabolites required for their growth and survival.

## Methods

### Clinical Study Selection

The cohort for this study consisted of patients with metastatic breast cancer and varying status of estrogen-receptor positive, progesterone receptor positive, or HER2 positive. The patients selected were on a stable treatment for ≥ 6 weeks and had no planned upcoming changes to their cancer therapy. The patients were screened to meet the requirements needed to participate, such as exclusions for patients with excessive alcohol intake, smoking, use of certain medications such as blood thinners, or any alteration that could be a confounding factor, such as recent surgery or recent implementation of the vegan diet. The screening for the participants consisted of a group information session regarding the dietary intervention and an individual visit to review medical history and confirm eligibility for the study. If the participants did not complete the screening or meet eligibility requirements, they were excluded from the study. Participants were randomized into two cohorts with blocks of 4 for the clinical study. The participants on the ad libitum whole food plant-based diet (n=21) were given three meals and a side dish daily for a total of 8 weeks. Four intervention subjects were not included in this analysis. One forgot to complete their final three-day food record, another was withdrawn from the study by investigators early in their participation due to the pandemic shutdown, two were deemed poorly compliant with the study diet, one fully non-compliant, by pre-defined criteria, and one only borderline compliant with high intake of animal-based foods. The control group (n=11) were asked to maintain their regular diet and were not included in this analysis. Participants attended weekly visits with study physicians and completed assessments at baseline, week 4, and week 8.

### Ad libtum Whole Food Plant-based Diet

The whole food plant-based diet excluded animal products and added fats and oils. The diet predominately consisted of vegetables, fruits, nuts and seeds, legumes, whole grains, and potatoes with modest amounts of soy and small amounts of sweeteners. Patients could consume their own foods in addition to, or in place of, the provided meals if their foods were ‘on plan.’ Participants were asked to consume as much as they wanted and eat as frequently as they wanted to feel satiated. Assessment of the patient’s dietary intake consisted of a 3-day food record at baseline and again at 8 weeks and three unscheduled 24-hour phone calls from a dietitian. The 3-day record consisted of two weekdays and one weekend day. The records were analyzed using the Nutrition Data System for Research (NDSR) version 2017 (Nutrition Coordinating Center, University of Minnesota, Minneapolis, MN). To calculate the percent difference in dietary components (i.e., consumption, calories, fat, carbohydrates, total protein, animal protein, vegetable protein, fiber, and amino acids), an average value was calculated based on the 3 daily food records at baseline. For the percent difference at baseline, the difference between the baseline value and the baseline average was divided by the baseline average and multiplied by 100%. For the percent difference at the 8-week follow-up, the difference between the baseline value and the baseline average was divided by the baseline average and multiplied by 100%.

### Serum Analysis

Serum samples for this analysis were collected at baseline and week 8. Serum samples (20 µl) were added to an extraction solution (1000 µl; 80% methanol with internal standards), vortexed, and centrifuged at max speed for 5 minutes. The supernatant (900 µl) was transferred to a new microcentrifuge tube and stored at -80 C. For metabolite preparation, 900 µL of metabolite extracts were dried with N_2_ gas and reconstituted in 90 µL of 50% acetonitrile (A955, Fisher Scientific).

For LC-MS analysis, metabolite extracts were analyzed by high-resolution mass spectrometry with an Orbitrap Exploris 240 (Thermo) coupled to a Vanquish Flex liquid chromatography system (Thermo). 2 µL of samples were injected into a Waters XBridge XP BEH Amide column (150 mm length × 2.1 mm id, 2.5 µm particle size) maintained at 25C, with a Waters XBridge XP VanGuard BEH Amide (5 mm × 2.1 mm id, 2.5 µm particle size) guard column. Mobile phase A was 100% LC-MS grade H_2_O with 20 mM ammonium formate and 0.1% formic acid. Mobile phase B was 90% acetonitrile with 20 mM ammonium formate and 0.1% formic acid. The gradient was 0_Jminutes, 100% B; 2_Jminutes, 100% B; 3_Jminutes, 90% B; 5_Jminutes, 90% B; 6_Jminutes, 85% B; 7_Jminutes, 85% B; 8_Jminutes, 75% B; 9_Jminutes, 75% B; 10_Jminutes, 55% B; 12_Jminutes, 55% B; 13_Jminutes, 35%, 20_Jminutes, 35% B; 20.1_Jminutes, 35% B; 20.6_Jminutes, 100% B; 22.2 minutes, 100% all at a flow rate of 150 μl min−1, followed by 22.7_Jminutes, 100% B; 27.9_Jminutes, 100% B at a flow rate of 300 μl min−1, and finally 28_Jminutes, 100% B at flow rate of 150 μl min−1, for a total length of 28 minutes. The H-ESI source was operated in positive mode at spray voltage 3500 with the following parameters: sheath gas 35 au, aux gas 7 au, sweep gas 0 au, ion transfer tube temperature 320 C, vaporizer temperature 275 C, mass range 70 to 800 m/z, full scan MS1 mass resolution of 120,000 FWHM, RF lens at 70%, and standard automatic gain control (AGC). LC-MS data were analyzed by El-Maven software(30), and compounds were identified by matching to LC-MS method-specific retention time values of external standards. Samples were run in two batches. To calculate relative abundance, peak intensities for baseline and 8-week follow-up samples were divided by the average peak intensity at baseline within each batch.

## Results

### A whole food, plant-based diet decreases the consumption of calories, fat, and protein in patients with metastatic breast cancer

A whole food, plant-based diet is reported to contain less protein than traditional animal-based diets (i.e., Western diets)(28, 29, 31). Patients with metastatic breast cancer enrolled in clinical trial NCT03045289 were provided an ad-libitum whole food, plant-based diet for 8 weeks(32, 33). Notably, participants were very adherent to the diet and reported positive outcomes regarding perceived cognitive function, emotional well-being, fatigue, and hunger(32). Participants documented their diets using three-day food records and unannounced 24-hour food recalls, and serum was collected at baseline and 8 weeks (Figure 1a). Analysis of food records and recalls found that participants consumed more total mass (including liquids) but less calories and fat (Figure 1b-1d). The consumption of carbohydrates by participants trended towards an increase but was non-significant (Figure 1e). These findings demonstrate that the adoption of a whole food, plant-based diet lowers the intake of calories and fat in patients with metastatic breast cancer. Since both caloric and fat consumption are predictors of tumor growth and progression(24–26), this suggests that a whole food, plant-based diet can potentially benefit patients.

**Figure 1.**
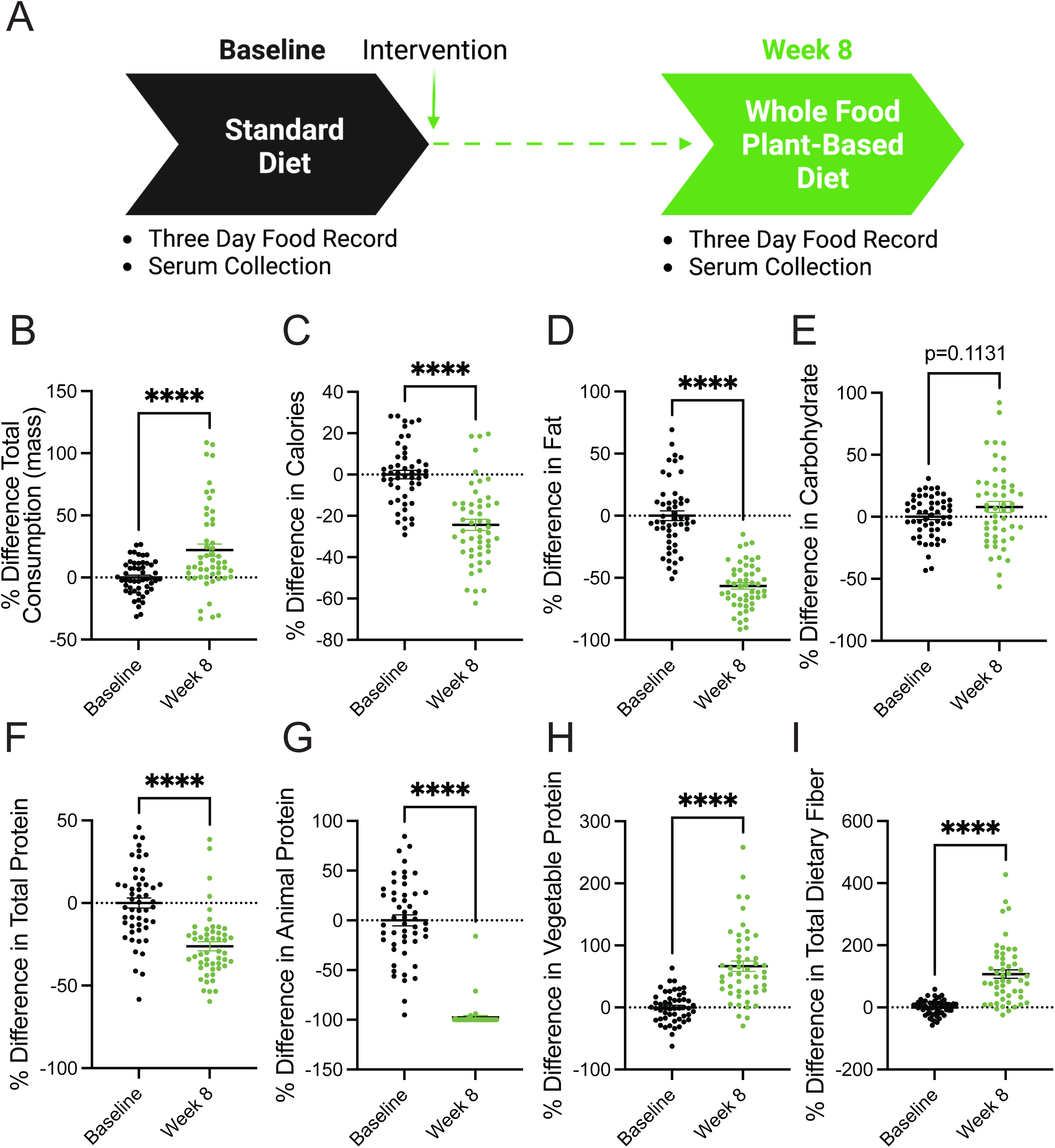
A whole food, plant-based diet decreases the consumption of calories, fat, and protein in patients with metastatic breast cancer. (A) Patients with metastatic breast cancer adhered to a whole food plant-based diet for 8 weeks. Two 3-day food records and three unannounced 24-hour recalls documented their dietary intake. Serum was collected at baseline and 8 weeks. (B-I) The percent difference between baseline and 8-week follow-up was measured in participants’ (n=51; 17 participants each with 3 daily food records at baseline and 8-week follow-up) consumption of total mass (B), calories (C), fat (D), carbohydrates (E), total protein (F), animal protein (G), vegetable protein (H), and fiber (I) following the adoption of a whole food, plant-based diet. An unpaired t test was used to determine statistical significance. ** P value < 0.01, **** P value < 0.0001.

In pre-clinical models, plant-based diets are reported to lower the consumption of total protein (i.e., amino acids)(29). In line with these findings, we found that participants adhering to a whole food, plant-based diet consumed significantly less protein (Figure 1f). Further, there was a near-complete substitution of animal protein with vegetable protein (Figure 1g-1h). These findings demonstrate that the adoption of whole food, plant-based diet is a clear approach to lowering the consumption of amino acids in patients with cancer. Since cancers have an elevated demand for amino acids(1, 2), the consumption of a whole food, plant-based diet is potentially an opportunity to deprive cancers of these amino acids.

Increased dietary fiber can support improved responses to tumor immunotherapy(23), and participants were found to consume increased amounts of total dietary fiber (Figure 1i). This suggests that, along with decreasing the consumption of pro-tumorigenic metabolites (i.e., fat and protein), adopting a whole food, plant-based diet increases the consumption of metabolites (i.e., fiber) that support anti-cancer therapies.

### Consumption of a whole food, plant-based diet decreases serum levels of cholesterol, insulin, and IGF

A diet high in whole foods includes those lower in energy density, thus leading to lower calorie intake(27, 32, 34). Additionally, the consumption of whole food results in increased calories lost in feces (i.e., lower metabolizable energy)(35), which could support weight loss. Further, increased consumption of vegetables, such as with the dietary approaches to stop hypertension (DASH) diet, is well-established to be beneficial towards improved metrics of cardiovascular health (i.e., lower levels of cholesterol, HDL, and LDL)(36, 37). In line with the decreased caloric intake (Figure 1c), metastatic breast cancer patients consuming a whole food, plant-based diet showed decreases in body mass index (BMI) and weight (Figure 2a-2b). While no differences in fasting serum triglycerides were observed (Figure 2c), patients had lower levels of serum cholesterol, HDL, and LDL (Figure 2d-2f). These results demonstrate that the adoption of a whole food, plant-based diet by patients with cancer supports weight loss and lowers prognostic markers of cardiovascular disease.

**Figure 2.**
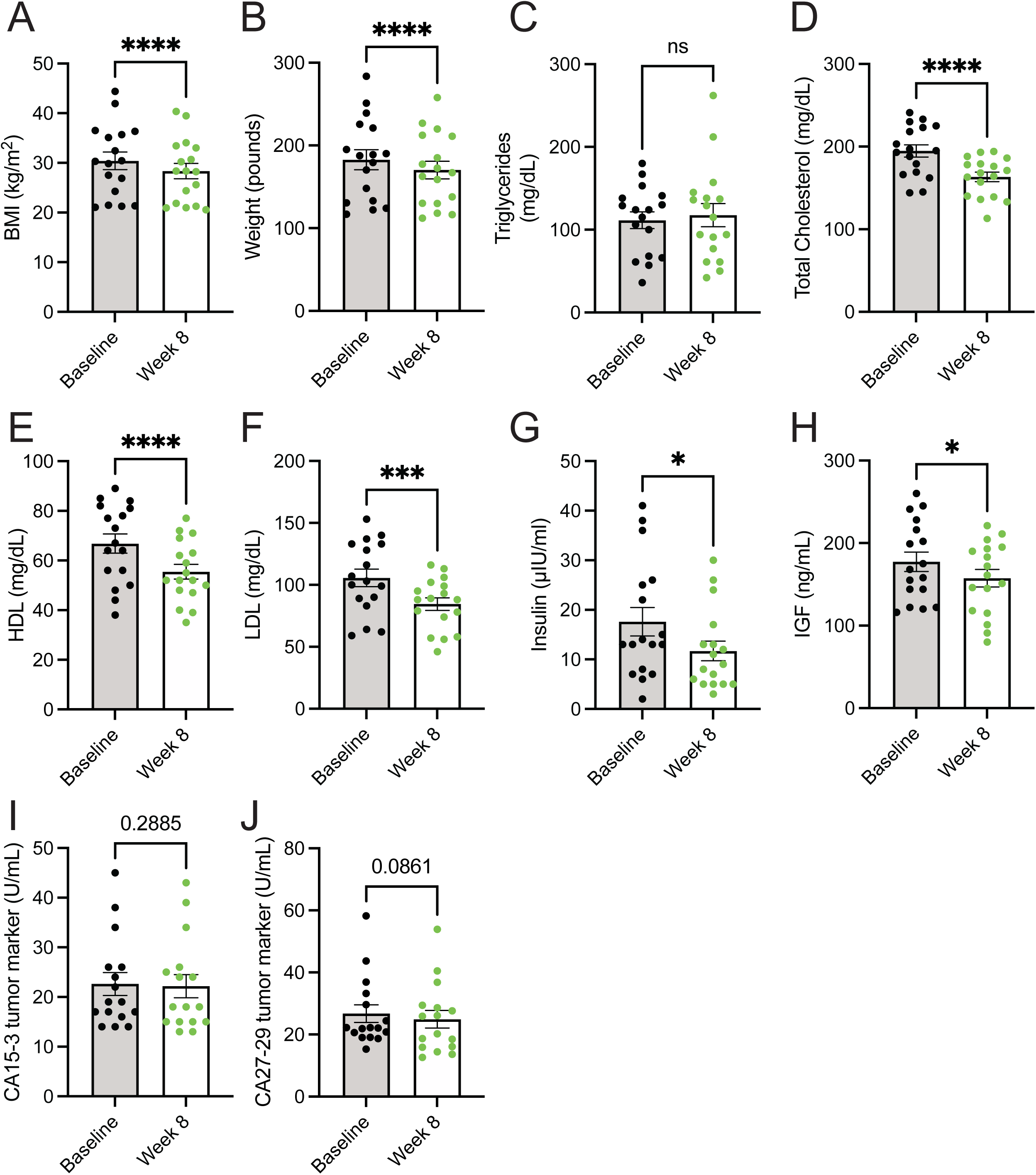
Consumption of a whole food, plant-based diet decreases serum levels of cholesterol, insulin, and IGF. (A-J) Body mass index (BMI) (A) and weight (B), and serum levels of triglycerides (C), total cholesterol (D), HDL (E), LDL (F), insulin (G), IGF (H), CA15-3 (I), and CA27-29 (J) were measured in patients at baseline and 8-week following the adoption of a whole food, plant-based diet. n = 17 for (A-H) and n = 16 for (I-J). A paired t test was used to determine statistical significance. ns = not significant, * P value < 0.05, *** P value < 0.001, **** P value < 0.0001.

Numerous dietary factors(37–40), including protein intake(41–46), are reported to impact the abundance of growth hormones, such as insulin and insulin-like growth factor (IGF). These growth factors can signal molecular pathways to drive cancer growth and survival(47, 48). Thus, limiting dietary protein would potentially lower the growth factors involved in glucose metabolism (i.e., insulin and IGF), impair cancer growth, and sensitize tumors with anti-cancer therapies, such as PI3K inhibitors(49). Less is known whether lowering dietary proteins through a plant-based diet can lower levels of these growth factors. We found that participants consuming a whole-food, plant-based diet had lower levels of insulin and IGF (Figure 2g-2h). Further, these reductions in insulin and IGF occurred not only in the absence of a decrease in dietary carbohydrates but in a non-significant but trending increase in dietary carbohydrates (Figure 1f). These results demonstrate that adopting a whole food, plant-based diet is a potential approach to lowering serum levels of tumor-promoting growth factors (i.e., insulin and IGF).

Limiting the dietary intake of total protein(14–17) or specific amino acids(18–21) has been shown to impede tumor growth in preclinical cancer models. While a change in tumor volume was not a primary endpoint of the 8-week clinical trial(32), we sought to examine if there were any potential changes in circulating tumor markers following the adoption of the diet. Serum markers CA15-3 and CA27-29, which correlate with breast tumor burden(50), showed non-significant but trending decreases in patients following the implementation of a whole food, plant-based diet (Figure 2i-2j). Future clinical trials with extended time frames (i.e., greater than 8 weeks) are required to better understand how a whole food, plant-based diet impacts tumor progression in patients.

### A whole food, plant-based diet reduces the levels of serum amino acids

Plant-based diets are reported to contain lower amounts of protein compared to animal-based diets(28). Notably, equivalent reductions are described to occur across all amino acid species with a plant-based diet(29). In line with these studies, we found that patients on a whole food, plant-based diet consumed fewer amino acids, which occurred to both essential and non-essential amino acids (Figure 3a-3c). However, a broad range of reductions in amino acid species was observed, some with minimal and non-significant reductions (i.e., arginine and aspartate) and others with dramatic reductions (i.e., methionine and proline). Further, due to the technical approaches for quantification of amino acids in foods, the levels of glutamine and asparagine in the diets were not determined. These findings show that the consumption of a whole food, plant-based diet by patients with metastatic breast cancer significantly lowers their intake of essential and non-essential amino acids.

**Figure 3.**
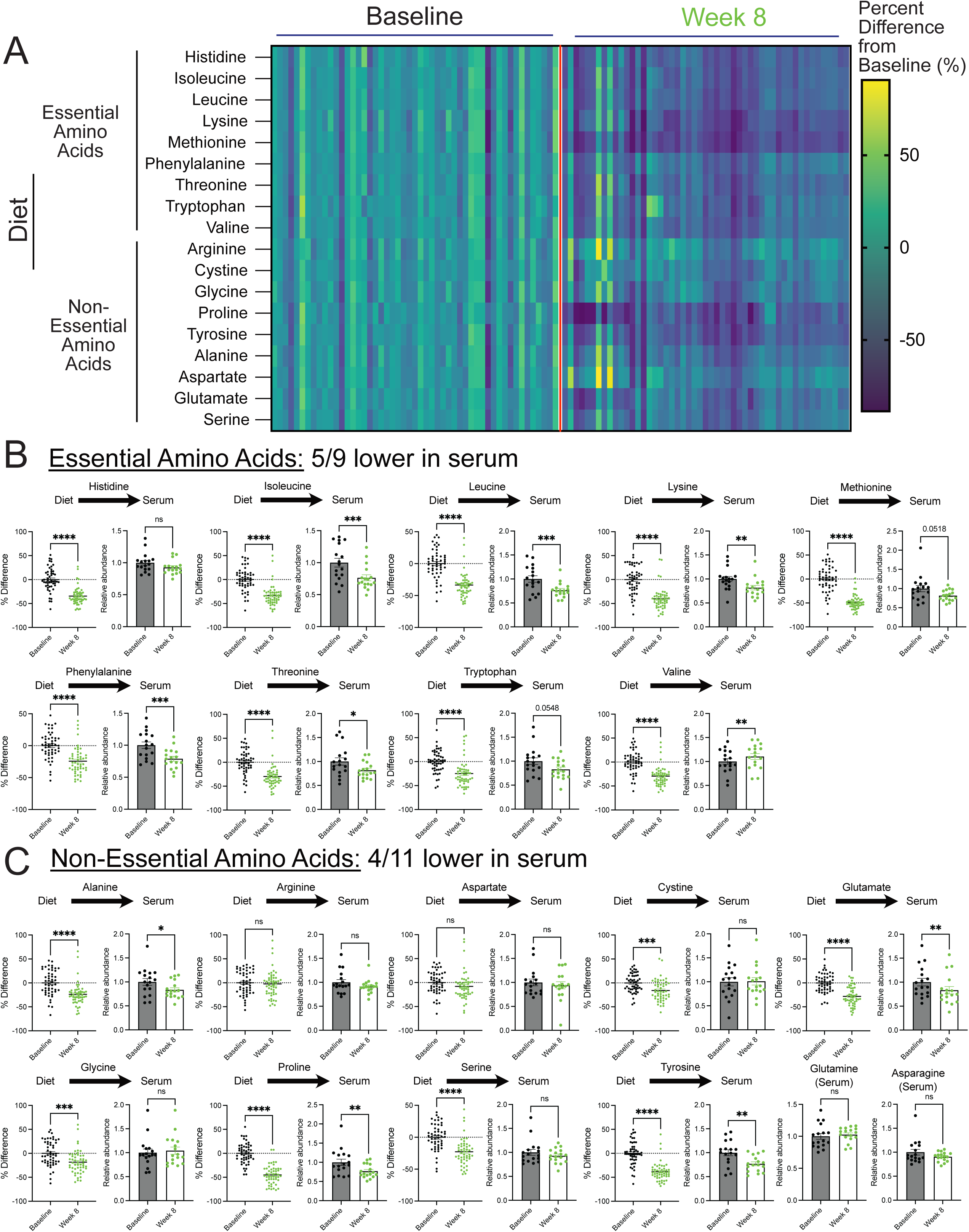
A whole food, plant-based diet reduces the levels of serum amino acids. (A-C) The percent difference in amino acid consumption between average and 8-week follow-up and the relative abundance of essential (B) and non-essential (C) amino acids in the serum of patients at baseline and 8-week follow-up. An unpaired t test was used to determine the statistical significance of the percent difference in dietary consumption of amino acids (n=51; 17 participants each with 3 daily food records at baseline and 8-week follow-up). A paired t test was used to determine the statistical significance of the relative abundance in serum levels of amino acids (n = 17). ns = not significant, * P value < 0.05, ** P value < 0.01, *** P value < 0.001, **** P value < 0.0001.

Essential amino acid levels are suggested to be maintained through dietary intake. Thus, we examined whether lowered consumption of essential amino acids in the diet translated to lower levels in the serum of patients. Interestingly, while we found reductions, this was not the case for every essential amino acid, as only 5/9 were significantly lowered (i.e., isoleucine, leucine, lysine, phenylalanine, and threonine) (Figure 3b). Certain amino acids (i.e., tryptophan and methionine) saw reductions but were not statistically significant. An outlier was valine, which had increased levels in the serum. These findings demonstrate that while essential amino acids are described to be solely maintained from dietary sources, reductions in the dietary intake of these amino acids do not predict reductions in the serum. Nonetheless, this shows that reducing essential amino acids in patients with cancer is clinically achievable and may contribute to slower tumor growth.

Unlike essential amino acids, the levels of non-essential amino acids are controlled by de novo synthesis and dietary intake. Thus, it was unclear if limiting non-essential acids would result in lower levels in patients. We found 4/11 non-essential amino acids (i.e., alanine, glutamate, proline, tyrosine) were lower in the serum of participants consuming a whole food, plant-based diet (Figure 3c). Some of these reductions in non-essential amino acids could be due to reductions in essential amino acids; however, it was unclear why certain amino acids were reduced and others weren’t (i.e., cystine, serine, glycine). Nonetheless, these results show that, along with reductions in serum levels of essential amino acids, the consumption of a whole food, plant-based diet leads to reductions in serum levels of non-essential amino acids in patients with cancer.

## Discussion

Limiting the dietary intake of amino acids has been shown to slow tumor growth in pre-clinical models of cancer. We show that consuming a whole food, plant-based diet lowers the dietary intake and circulating levels of amino acids in patients with cancer. These studies provide a framework for clinically exploring the efficacy of lowering amino acids on slower tumor growth. Notably, significant efforts have been placed on limiting non-essential amino acids from the diet (i.e., serine and glycine)(51), but few studies have explored the possibility of limiting essential amino acids in patients, potentially due to the perception that limiting essential acids could be harmful. We find lowering both essential and non-essential amino acids in patients with cancer does not have a harmful impact; instead, participants reported a positive impact on several reported outcomes, including perceived cognitive function, emotional well-being, and fatigue(32). However, one consideration is that, compared to preclinical studies(18–21), this diet lowered these amino acids rather than eliminated them. Future clinical trials, with extended time frames, are required to examine if lowering these amino acids impacts tumor growth in patients.

Lowering circulating growth factors (i.e., insulin and IGF) is a therapeutic approach to improving anti-cancer drug efficacy(49), and dietary protein abundance can drive the production of these growth factors(44, 45). We found that adopting a whole food, plant-based diet reduced the levels of insulin and IGF in participants. This occurred with a non-significant but trending increase in dietary carbohydrates. These findings suggest a clear approach to limiting these pro-tumor growth factors that do not require limiting the consumption of carbohydrates. Further, it suggests that combining this diet with anti-cancer therapies targeting these signaling pathways (i.e., PI3K inhibitors) would potentially act synergistically in slower tumor growth.

Essential amino acids cannot be synthesized and must be consumed from the diet. We found that a whole food, plant-based diet lowered the dietary intake of every essential amino acid. Surprisingly, this did not translate into a corresponding reduction in every essential amino acid in the serum of patients. Notably, levels of valine showed an increase in levels, which is counterintuitive. Further research, including a longer trial duration and larger sample size, is required to better understand how the levels of essential amino acids are regulated in the body and impacted by diet.

Numerous diets are suggested to deprive tumors of nutrients and improve outcomes in patients with cancer(52). However, the probability of patients adopting these diets is more limited. We found that switching from an animal-based diet (i.e., Western diet) to a whole food, plant-based diet was feasible without any limitations to patients’ quality of life. On the contrary, participants reported positive outcomes from the diet. A potential explanation is that this diet was provided ad libitum, without any limitation in food intake, and participants consumed more total food mass. Further, since this diet requires no special formulation (i.e., protein shakes), it is a highly tractable approach. Overall, this data suggests that the consumption of a whole food, plant-based diet is a clear method of lowering circulating amino acids in patients with cancer and potentially an approach to deprive tumors of nutrients required for their growth and survival.

## Data Availability

All data produced in the present study are available upon reasonable request to the authors.

## Abbreviations

IGF: insulin-like growth factor
HER2: human epidermal growth factor receptor 2
NDSR: nutrition data system for research
LC-MS: liquid chromatography-mass spectrometry
DASH: dietary approaches to stop hypertension
BMI: body mass index
HDL: high-density lipoprotein
LDL: low-density lipoprotein
PI3K: phosphoinositide 3-kinase
CA15-3: cancer antigen 15-3
CA27-29: cancer antigen 27-29

## Declarations

### Ethics approval and consent to participate

This clinical study occurred at the University of Rochester Medical Center and complied with recognized ethical guidelines, including the U.S. Common Rule. The study protocol was approved by the University of Rochester Research Subject Review Board (ClinicalTrials.gov identifier: NCT03045289), and written informed consent was obtained from all participants.

### Consent for publication

Not applicable.

### Availability of data and materials

No datasets were generated during the current study.

### Authors’ contributions

T.Q.S. and I.S.H. initiated the study and conceived the project, designed experiments, interpreted results, and wrote the manuscript. B.S. and J.M. performed metabolite analysis. L.M.B., N.W., T.M.C., and E.K.C. conducted the clinical trial and sample acquisition. E.T.T., B.J.A., and L.J.P. supported project administration and provided expert comments.

## Acknowledgments

We would like to thank the Metabolomics Resource and the Center for Advanced Research Technologies (CART) at URMC. The schematic in Figure 1 was created by BioRender.

